# COVID-19 and first trimester spontaneous abortion: a case-control study of 225 pregnant patients

**DOI:** 10.1101/2020.06.19.20135749

**Authors:** Stefano Cosma, Andrea Carosso, Jessica Cusato, Fulvio Borella, Marco Carosso, Marialuisa Bovetti, Claudia Filippini, Antonio D’Avolio, Valeria Ghisetti, Giovanni Di Perri, Chiara Benedetto

**Author notes:** **Correspondence address:** Stefano Cosma, Gynecology and Obstetrics 1, Department of Surgical Sciences, City of Health and Science, University of Turin, Via Ventimiglia 3, 10126, Turin, Italy, /+39 011 3131711. Joint first authors (Stefano Cosma, Andrea Carosso).

## Abstract

**Background:** Evidence for the impact of COVID-19 during the second and the third trimester of pregnancy is limited to a relatively small series, while data on the first trimester are scant. With this study we evaluated COVID-19 infection as a risk factor for spontaneous abortion in first trimester of pregnancy.

**Methods:** Between February 22 and May 21, 2020, we conducted a case-control study at S. Anna Hospital, Turin, among first trimester pregnant women, paired for last menstruation. The cumulative incidence of COVID-19 was compared between women with spontaneous abortion (case group, n=100) and those with ongoing pregnancy (control group, n=125). Current or past infection was determined by detection of SARS-CoV-2 from nasopharingeal swab and SARS-CoV-2 IgG/IgM antibodies in blood sample. Patient demographics, COVID-19-related symptoms, and the main risk factors for abortion were collected.

**Findings:** Twenty-three (10.2%) of the 225 women tested positive for COVID-19 infection. There was no difference in the cumulative incidence of COVID-19 between the cases (11/100, 11%) and the controls (12/125, 9.6%) (p=0.73). Logistic regression analysis confirmed that COVID-19 was not an independent predictor of abortion (1.28 confidence interval 0.53-3.08).

**Interpretation:** COVID-19 infection during the first trimester of pregnancy does not appear to predispose to abortion; its cumulative incidence did not differ from that of women with ongoing pregnancy.

## Introduction

The World Health Organization (WHO) named the new coronavirus (SARS-CoV-2) disease coronavirus disease-19 (COVID-19) and declared it a pandemic. Coronaviruses are enveloped, non-segmented positive-sense RNA usually responsible for mild illness such as the common cold in adults and children.^1^ But in the last decade, coronaviruses have caused two important epidemics: the severe acute respiratory syndrome (SARS) and the Middle East respiratory syndrome (MERS). COVID-19 was first reported in Wuhan (China) in December 2019 followed by outbreaks across the world.^2^ The first cases of COVID-19 in Italy were confirmed in January 2020, with a rapid rise in the number of cases in northern Italy starting in late February.

Despite the rapidly growing number of cases worldwide, data on COVID-19 during pregnancy remain limited, being derived mainly from small sample studies.^3–8^ A systematic review of published reports on coronaviruses (COVID-19, SARS, MERS) reported higher rates of preterm birth, preeclampsia, cesarean section, and perinatal death.^9^ The lack of data on abortion due to COVID-19 during the first trimester precludes extrapolation of conclusive evidence for the effects of infection during early pregnancy. The paucity of reliable data has aroused concern in patients, while the disinformation reported by media may lead pregnant women to embrace dramatic choices such as voluntary abortion.^10^ The wide of clinical expression, the high rate of asymptomatic forms, the poor accuracy of nasopharyngeal swab testing and its limited availability have been the main barriers to gaining a real understanding of the prevalence of the infection and its impact on pregnancy. In this complex scenario, the development of serological tests for the detection of SARS-CoV-2 IgG and IgM could be useful to identify pregnant patients who were infected during early pregnancy. While the quantity and quality of data on test performance are still limited, the level of accuracy has been reportedly moderate/good, so that patients infected by SARS-CoV-2 can be traced.^11^

The aim of the present study was to evaluate the impact of COVID-19 on first trimester spontaneous abortion by comparing the cumulative incidence of SARS-CoV-2 infection in a cohort of women who experienced early abortion and that of women with ongoing pregnancy at 12 weeks of gestational age.

## Materials and methods

Women who had been referred to our Hospital for first trimester spontaneous abortion care between February 22 and May 21, 2020 were contacted and enrolled (case group). Women 12 weeks pregnant admitted to our Hospital for fetal nuchal translucency between April 16 and May 21, 2020 were the control group. The first reported case of COVID-19 infection in Piedmont was dated February 22, 2020. To exclude the possibility of COVID-19 seroconversion before pregnancy, only women with last menstruation before that date were considered eligible for inclusion (Fig. 1). This criterion allowed us to define seropositivity in the case group as a seroconversion that had occurred during pregnancy. Blood tests were performed for the detection of IgG/IgM non neutralizing antibodies against SARS-CoV-2 and reverse transcriptase-polymerase chain reaction (RT-PCR) assays on nasopharingeal swabs. Patients testing positive at least one test were also tested for the determination of specific neutralizing antibodies. Blood samples were centrifuged at 3000 rpm for 5 min to separate serum and analyzed the same day of collection.

**Figure 1:**
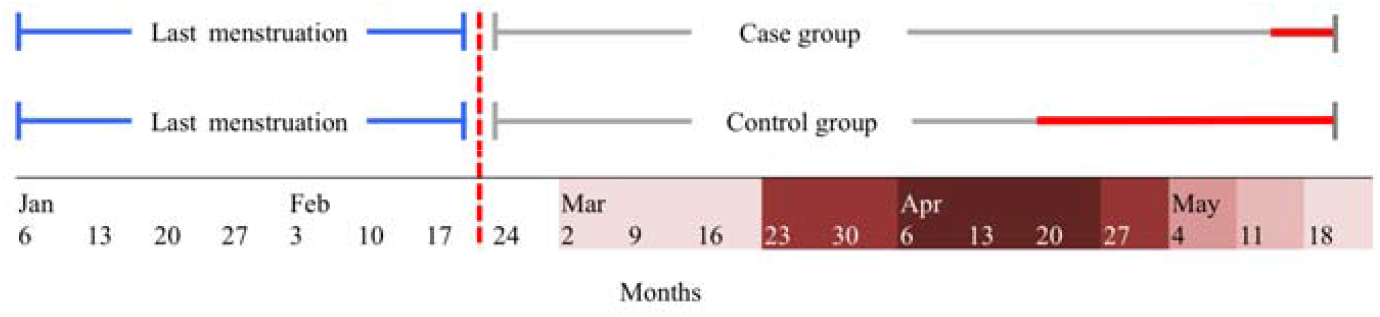
Inclusion criteria and time of serological and molecular sampling in the case and the control group. Blue line: time range for last menstruation inclusion; dotted red line: first reported case of COVID-19 in Piedmont, Italy; red line: time of sera and nasopharyngeal swab sample collection 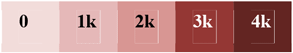 COVID-19 outbreak cases in Piedmont Region: weekly case increase

A rapid automated fluorescent lateral flow CE-approved immunoassay (AFIAS™ COVID-19, Boditech, Gang-won-do, Korea) was used for qualitative and semi-quantitative detection of IgG/IgM non neutralizing antibodies against the spike (S) and nucleocapsid (N) viral proteins; semi-quantitative results are expressed as the cut-off index (COI) in which a COI > 1.1 indicates a positive result. Chemiluminescence CE-approved immunoassay (CLIA) technology was used for the semi-quantitative determination of anti-S1 and anti-S2 specific IgG neutralizing antibodies to SARS-CoV-2 (Liaison® SARS-CoV-2 S1/S2 IgG, Diasorin, Saluggia, Italy): the antibody concentration is expressed as arbitrary units (AU/mL) and grades the results as positive when ≥ 15 AU/mL. Viral RNA extraction from the swab was performed on a MagNA Pure compact instrument (Roche, Mannheim, Germany) and analyzed using a RT-PCR assay (CFX-96, Bio-Rad, Milan, Italy) with the Liferiver Novel Coronavirus 2019-nCov real-time RT-PCR kit protocol, targeting genes N, E, and ORF1ab (Liferiver Bio-Tech, San Diego, CA, USA).

Sample size calculation was not possible because the expected prevalence of disease was unknown at the time of population enrollment and further recruitment beyond May 21 would have precluded the eligibility criterion for last menstruation.

Demographics, COVID-19-related symptoms, and data on exposure to possible risk factors for abortion were collected by interview. The study was approved by the Institutional Review Board of the City of Health and Science of Turin (Reference number: 00171/2020). Written, informed consent was obtained from all participants. The results for quantitative variables are expressed as the mean ± standard deviation (SD) and qualitative categorical variables are expressed as frequency and percentages. Comparison of quantitative variables was performed using the t-test or Wilcoxon-Mann–Whitney test based on normal or not distribution, respectively. Qualitative variables were compared using the chi-square test or Fisher’s exact test, as appropriate. When basic patient characteristics were present as confounding factors, regression analysis was performed to assess the relationship between COVID-19 infection and spontaneous abortion. Results are expressed as odds ratio (95% confidence interval [CI]). Statistical analyses were performed using SAS software ver. 9.4 for Windows (SAS Institute, Carey, NC, USA).

## Role of funding source

The funders of the study had no role in study design, data collection, data analysis, data interpretation, or writing of the report. The corresponding author had full access to all the data in the study and had final responsibility for the decision to submit for publication.

## Results

A total of 225 women at first trimester of pregnancy, attending our Institute were included in the study. One hundred women in the case group and 125 women in the control group were enrolled. The patient adhesion rate was 87% (100/115) and 88% (125/142), respectively. Table 1 presents the patients’ characteristics at baseline; except for age, there were no statistically significant differences in demographics or risk factors for abortion between the two groups. Twenty-three of the 225 women tested for anti-SARS-CoV-2 IgG and IgM antibodies were found to be seropositive or their nasopharyngeal swab tested positive for COVID-19, yielding an overall cumulative incidence of 10.2% in the first trimester. There was no significant difference in the cumulative incidence of COVID-19 between the case patients (11/100, 11%) and the controls (12/125, 9.6%) (p=0.73).

**Table 1.** Baseline characteristics, clinical findings, and COVID-19 cumulative incidence in case and control groups.

| Clinical findings |  | Case<br>N=100 | Control<br>N=125 | p-value |
| --- | --- | --- | --- | --- |
| | | No. (%) or mean (±SD) | No. (%) or mean (±SD) | |
| Age | | 35.5 (±4.7) | 33.7 (±4.7) | 0.001 |
| BMI prior to pregnancy, Kg/m<sup>2</sup> | | 25.5 (±4.3) | 22.6 (±4.1) | 0.11 |
| Pregnancy | 0 | 51 (51) | 77 (61.6) | 0.34 |
|  | 1 | 40 (40) | 37 (29.6) |  |
|  | 2 | 7 (7) | 9 (7.2) |  |
|  | 3 | 1 (1) | 2 (1.6) |  |
|  | 5 | 1 (1) | 0 (0) |  |
| Previous abortion | 0 | 66 (66) | 94 (75.2) | 0.11 |
|  | 1 | 27 (27) | 21 (16.8) |  |
|  | 2 | 6 (6) | 7 (5.6) |  |
|  | 3 | 0 (0) | 3 (2.4) |  |
|  | 6 | 1 (1) | 0 (0) |  |
| ART therapy |  | 7(7) | 12(9.6) | 0.48 |
| Smoking history |  | 22 (22) | 16 (12.8) | 0.06 |
| Thyroid disease |  | 10 (10) | 11 (8.8) | 0.75 |
| Autoimmune diseases |  | 8 (8) | 4 (3.2) | 0.11 |
| Thrombophilia |  | 5 (5) | 5 (4) | 0.75 |
| Uncontrolled DM |  | 0 | 0 | >0.99 |
| Uterine abnormalities |  | 8 (8) | 9 (7.2) | 0.82 |
| COVID-19 disease |  | 11 (11) | 12 (9.6) | 0.73 |

Ab, antibodies; ART, assisted reproductive technique; DM, diabetes mellitus

The age variable was entered into logistic regression analysis to evaluate COVID-19 infection in relation to confounders. There was no difference in the odd of being infected with SARS-CoV-2 between the two groups, indicating that COVID-19 infection was not an independent predictor of abortion (1.282, CI 0.53-3.08).

In the case group, 5/11 (45.4%), 3/11 (27.2%), and 1/11 (9%) were positive for SARS-CoV-2 IgG, SARS-CoV-2 IgM, or both SARS-CoV-2 IgG and IgM, respectively; RT-PCR of the nasopharingeal swab resulted positive in 2/11 (18%) (Table 2). In the control group, 7/12 (58.3%), 3/12 (25%), and 2/12 (16.6%) were positive for SARS-CoV-2 IgG, SARS-CoV-2 IgM, or both SARS-CoV-2 IgG and IgM, respectively; RT-PCR of the nasopharingeal swab resulted positive in 5/12 (41.7%) (Table 3). No difference in positivity for IgG neutralizing antibodies was found between the case (6/11, 54.5%) and the control group (5/12, 41.7%) (p=0.53) (Table 1). There was no statistically significant difference between the two groups for average antibody titer, both non neutralizing (21.3 vs. 18.3 COI; p=0.42) and neutralizing antibodies (39.9 vs 46.9 AU/ml; p=0.69).

**Table 2.** Antibody levels and SARS-Cov-2 detection in sera and nasopharyngeal swab samples from patients with abortion.

| Diagnostic Test | Positive result | Patient |  |  |  |  |  |  |  |  |  |  |
| --- | --- | --- | --- | --- | --- | --- | --- | --- | --- | --- | --- | --- |
|  |  | 1 | 2 | 3 | 4 | 5 | 6 | 7 | 8 | 9 | 10 | 11 |
| Anti-NP IgM | COI>1.1 | <1.1 | 2.11 | <1.1 | 1.9 | <1.1 | <1.1 | <1.1 | <1.1 | 2.6 | <1.1 | 2.9 |
| Anti-NP IgG | COI>1.1 | <1.1 | 18.9 | <1.1 | <1.1 | 19.4 | <1.1 | 14.4 | 32.4 | <1.1 | 21.7 | <1.1 |
| Anti-RBD IgG | ≥15 AU/ml | <15 | 19.5 | <15 | <15 | 29.9 | 49.3 | 17.3 | 41 | <15 | 82.9 | <15 |
| NS |  | pos | Neg | pos | neg | neg | neg | neg | neg | neg | neg | neg |

NS, nasopharyngeal swab; NP, nucleoprotein; RBD, receptor-binding domain

**Table 3.** Antibody levels and SARS-CoV-2 detection in sera and nasopharyngeal swab samples from pregnant patients.

| Diagnostic Test | Positive result | Patient |  |  |  |  |  |  |  |  |  |  |  |
| --- | --- | --- | --- | --- | --- | --- | --- | --- | --- | --- | --- | --- | --- |
|  |  | 1 | 2 | 3 | 4 | 5 | 6 | 7 | 8 | 9 | 10 | 11 | 12 |
| Anti-NP IgM | COI>1.1 | <1.1 | <1.1 | <1.1 | 2.1 | 1.6 | 1.2 | 1.2 | <1.1 | 1.2 | <1.1 | <1.1 | <1.1 |
| Anti-NP IgG | COI>1.1 | 19.3 | 19.3 | 15.6 | <1.1 | <1.1 | 21 | <1.1 | 21.5 | 23.2 | 21.9 | 2.45 | 20.7 |
| Anti-RBD IgG | ≥15 AU/ml | <15 | <15 | <15 | <15 | <15 | 52.7 | <15 | 21.1 | 103 | 30.5 | <15 | 27.5 |
| NS |  | neg | pos | neg | neg | neg | pos | neg | pos | neg | pos | neg | pos |

NS, nasopharyngeal swab; NP, nucleoprotein; RBD, receptor-binding domain

Twelve of the COVID-19 patients reported previous symptoms (12/23, 52.2%) including fever (7/12, 58.3%), anosmia and ageusia (5/12, 41.7%), cough (5/12, 41.7%), arthralgia (4/12, 33.3%), and diarrhea (1/12, 8.3%); no pneumonia or Hospital admission due to COVID-19-related symptoms was recorded. No difference in the incidence of symptoms was noted between the case (4/11, 36.4%) and the control group (8/12, 66.6%) (p=0.14).

## Discussion

With this case-control study, we evaluated the impact of COVID-19 on first trimester spontaneous abortion in a cohort of pregnant women with SARS-CoV-2 infection confirmed by antibody testing or RT-PCR assay of nasopharyngeal swabs. The results show that the risk of first trimester abortion is not impacted by SARS-CoV-2 infection, also after being adjusted for age. To the best of our knowledge, this may be the largest cohort of Coronaviruses infection during early pregnancy published so far.

The course of COVID-19 varies widely: patients may remain asymptomatic or develop mild to severe symptoms leading to pneumonia, respiratory failure, and death.^12^ The non-negligible prevalence of infection in asymptomatic pregnant women reported elsewhere^8,13^ makes universal screening of all pregnant patients appear desirable. However, because international guidelines diverge on this issue, it is difficult to determine the real impact that COVID-19 could have on pregnancy, especially during the first weeks of gestation, which are usually managed with outpatient monitoring; in some cases, abortion may be considered even before an obstetric exam has been made.

Serologic tests, in conjunction with SARS-CoV-2 RT-PCR assay, may offer a more feasible opportunity to identify both active and past infections and to evaluate the real spread of SARS-CoV-2, to the point that some governments have suggested their use in large-scale population tracking.^14^ Determination of seroconversion in pregnant women could answer some concerns about unfavorable pregnancy outcomes, which are not otherwise resolvable.

One of the strengths of the present study is the enrollment of women with serologically confirmed COVID-19 by means of two different serological assays; the combined results of RT-PCR on nasopharyngeal swab samples is another major strength of the study. The high adhesion rate to the study protocol limited confounding factors such as population selection bias. Antibodies to COVID-19 were detected in about one out of ten pregnant patients in the cohort; this finding should be carefully interpreted, however, as it cannot be generalized because derived from a single center located in a region with a high incidence of COVID-19.

A major limitation of the study is that we were unable to accurately backdate the time of infection in women with spontaneous abortion. In the absence of an IgG avidity test, we evaluated the time elapsed between the abortion and the blood test for antibody detection. The profile of antibodies against SARS-CoV-2 in this cohort was comparable with previous findings. Seroconversion of IgG or IgM within 20 days after symptom onset has recently been reported.^15^ The median day of seroconversion for both IgG and IgM was 13 days with a synchronous or a discordant pattern. In light of this evidence, seroconversion during pregnancy could be excluded (or be controversial) only in one patient (no. 4, Fig. 2) in the case group. The detection of IgM antibodies at 66 days after abortion does not preclude that seroconversion might have occurred after the loss of pregnancy.

**Figure 2:**
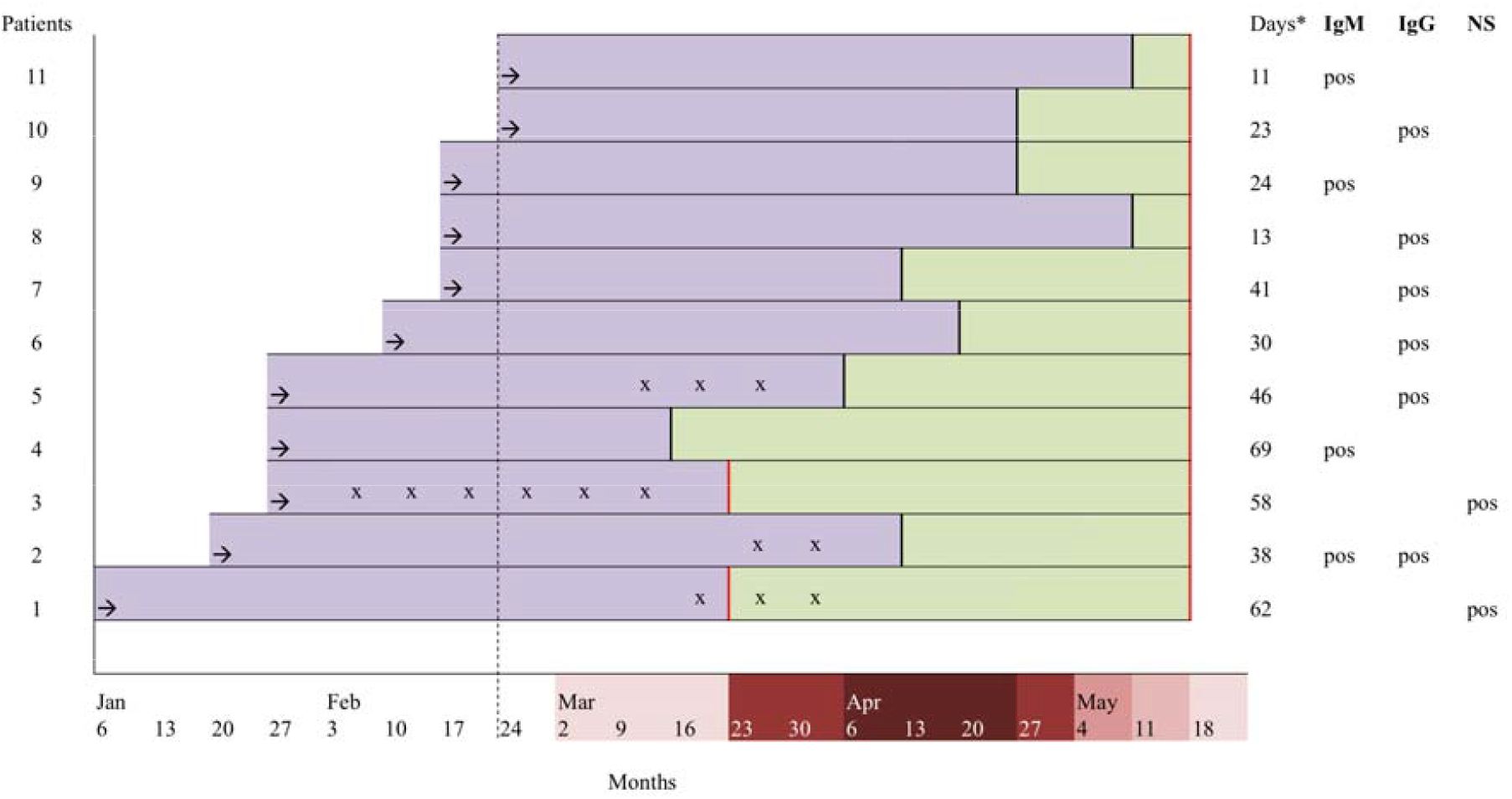
Patients with first trimester abortion: time elapsed between abortion care and diagnostic testing and seromolecular profiles. Black arrow: last menstruation; black vertical line: abortion Hospital care; dotted black line: first reported case of COVID-19 in Piedmont; NS: nasopharyngeal swab; rectangular green box: time elapsed between the abortion and diagnostic testing; rectangular violet box: pregnancy; red line: serological and/or molecular sampling; x: reported COVID-19-related symptoms; * days elapsed between the abortion and diagnostic testing 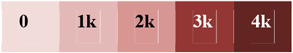 COVID-19 outbreak cases in Piedmont Region: weekly case increase

In view of future research addressing the issue on the relationship between COVID-19 and spontaneous abortion, it will be difficult for researchers to precisely define the timing of infection and the effective seroconversion during pregnancy. Inclusion criteria, together with the beginning of the study at pandemic outbreak, allowed us to fairly overcome this issue.

Concern is mounting about the impact of COVID-19 on pregnancy, possible vertical transmission,^16–18^ and unfavorable obstetric outcomes in particular. Reproductive medicine societies advised delaying the start of assisted reproductive treatments^19^ and guidelines on the prevention and control of COVID-19 among pregnant women have been issued.^20–22^ Currently, data on the impact of Coronaviruses on the first trimester of pregnancy are limited. Four of the seven patients who presented with SARS-CoV-1 infection during their first trimester had a spontaneous abortion, likely the result of the hypoxia caused by SARS-CoV-1-related acute respiratory distress.^23^ Furthermore, one case of a woman with MERS during the first trimester has been reported. She was asymptomatic and went on to have a term delivery.^24^ As for SARS-CoV-2, a single abortion during the second trimester of pregnancy in a woman with COVID-19 was probably related to placental infection.^25^ Another study reported the first visualization by electron microscopy of the SARS-CoV-2 invading syncytiotrophoblasts in the placental villi.^26^ This evidence could suggest a potential impact of SARS-CoV-2 on spontaneous abortion. However, our study findings may reduce concerns in patients during the first trimester of pregnancy. In the present cohort of women who experienced an abortion during the first trimester the serological prevalence of antibodies was similar to that in the women with ongoing pregnancies. Furthermore, although viral infection at this stage could potentially affect embryogenesis and organ development, there is still no evidence for the intrauterine transmission of SARS-CoV-2.

Despite these reassuring data, pregnancies in women with COVID-19 can still have an unfavorable obstetric outcome: inflammatory involvement of the placenta^27^ can be associated with preterm delivery.^28^ Moreover, physiologic maternal adaptations to pregnancy predispose pregnant women to a more severe course of pneumonia, with subsequent higher maternal and fetal morbidity and mortality.^29^ In this cohort, however, few patients were symptomatic and not more numerous in the case group. Severe disease was never observed. The lower incidence of severe manifestations during the first trimester could be explained by the minimal alteration in respiratory dynamics during this phase of pregnancy. In conclusion, our study provides reassuring findings for women who intend to become pregnant during the SARS-CoV-2 pandemic or who became infected during their first trimester of pregnancy. COVID-19 appears to have a favorable maternal course at the beginning of pregnancy, consistent with what has been observed during the third trimester when the clinical characteristics of COVID-19-positive pregnant women were similar to those found in women from the general population.^30^ More importantly, no significant difference in the early abortion rate was observed. Long-term follow-up of ongoing pregnancies will respond to other doubts about the impact of COVID-19 in pregnant patients.

## Data Availability

The dataset generated during during the current study is available from the corresponding author on reasonable request.

## Authors’ contributions

S.C. and A.C. had roles in the study design, data interpretation, literature search, and writing the article. J.C, V.G., A. D., had roles in experiments, and data collection. C.F. had roles in the data analysis and interpretation of the data. F.B., M. C. and M. B. had roles in recruitment, data collection, and execution of the study. G. DP. and C.B. contribute to coordinate the study. All authors reviewed and approved the final version of the manuscript.

## Declaration of interest

The authors report no conflict of interest

## Acknowledgments

We thank the staff of the Laboratory of S. Anna Hospital for sample collection and storage.

## Notes

**Disclosure:** The authors report no conflict of interest

**Financial Support:** research university funds

### Competing Interest Statement

The authors have declared no competing interest.

### Funding Statement

Research university funds

### Author Declarations

The study was approved by the Institutional Review Board of the City of Health and Science of Torino (Reference number: 00171/2020)

